# Improving ART Retention Through Machine Learning–Guided Targeting of Interventions: A Monte Carlo Simulation Study in Lilongwe, Malawi

**DOI:** 10.64898/2026.06.28.26356788

**Authors:** Agness Thawani, Bennett Kankuzi, Jacqueline Huwa, Layout Gabriel, Evelyn Viola, Ethel Rambiki

## Abstract

Retention in antiretroviral therapy (ART) care remains a major challenge in high-burden settings such as Malawi, where substantial loss to follow up undermines treatment outcomes and long-term epidemic control. Although machine learning models can accurately identify patients at high risk of disengagement, there is limited evidence on how these predictions can be translated into improved retention outcomes in practice. This study addresses this gap by explicitly linking machine learning–based risk stratification to the targeted allocation of retention interventions, providing a framework for evaluating their expected impact on ART retention outcomes. We developed a patient-level Monte Carlo simulation model that integrates individual predicted probabilities of loss to follow up from a validated Extreme Gradient Boosting model with intervention effect sizes derived from a meta-analysis of ART retention interventions conducted in sub-Saharan Africa. The study population included 1,705 ART patients receiving care at Lighthouse Trust clinics in Lilongwe, Malawi. Patients were stratified by predicted risk, and the highest-risk group (n = 512) was targeted for intervention. Six interventions were evaluated, including Expert Client support, psychosocial support, two-way text messaging, adherence clubs, community ART groups, and teen clubs, followed by subgroup-specific and combined approaches allocated based on predicted risk to reflect real-world programme implementation. The primary outcome was twelve-month ART retention, estimated over 5,000 simulation iterations. Subgroup and post-simulation analyses were conducted to assess heterogeneity in intervention response. Among patients classified as high risk (n = 512), baseline retention was 44.1%. Individual interventions improved retention to 52.7% with two-way texting (RR = 1.19; p < 0.001) and 55.0% with Expert Client support (RR = 1.25; p < 0.001). A combined intervention package produced substantially larger gains, increasing retention to 64.0% (RR = 1.45; p < 0.001), corresponding to an absolute improvement of 19.9 percentage points. Intervention effects varied across subgroups, with significant improvements observed among newly initiated patients (43.0% to 58.9%; RR = 1.37; p < 0.001) and clinically unstable patients (28.3% to 39.1%; RR = 1.38; p = 0.01), while effects among adolescents were more modest (34.3% to 45.6%; RR = 1.33; p = 0.03). Despite these improvements, 46% of high-risk patients remained hard to retain after receiving multiple interventions. In this subgroup, expected retention increased only marginally from approximately 0.15 at baseline to 0.20 after intervention, with particularly poor outcomes observed among patients who were virally unsuppressed, had depressive symptoms, or were younger. Machine learning–guided targeting of ART retention interventions can substantially improve retention outcomes, particularly when interventions are combined. However, a substantial subgroup of patients remains hard to reach and vulnerable to disengagement, indicating that existing strategies may be insufficient for individuals with complex clinical and psychosocial needs. This study contributes to knowledge by introducing an integrated framework that combines machine learning risk prediction, meta-analytic intervention effects, and patient-level Monte Carlo microsimulation to quantify twelve-month ART retention outcomes under risk-based targeting with subgroup-specific intervention allocation before real-world implementation. These findings highlight the potential of using individual risk to guide the delivery of retention interventions within routine ART programmes to enable more efficient, proactive, and patient-centered allocation of retention resources, while highlighting the need for more intensive and tailored approaches for individuals who remain hard to retain even after interventions.

**Author Summary:** Retaining people living with HIV (PLHIV) in long term care is essential for effective treatment and for reducing HIV transmission, yet many patients especially in high HIV burden and resource limited settings like Malawi disengage from antiretroviral therapy (ART). Current approaches, such as tracing patients after they miss clinic visits, tend to be reactive and can place a significant burden on already limited healthcare resources. While machine learning models can identify individuals at high risk of dropping out of care, they do not by themselves improve retention. Their value lies in helping health systems identify which patients should receive targeted support. However, there is limited evidence on how these predictions can be used to improve patient retention outcomes in practice.

In this study, we used machine learning predictions to identify individuals at high risk of dropping out of care and then simulated how retention could be improved by targeting different interventions to these patients. Using Clinical, behavioral and psychosocial data from two HIV clinics in Malawi, we estimated how different interventions, including peer support, psychosocial support, and two-way text messaging, perform when applied individually or in combination.

We found that targeting interventions based on predicted risk can substantially improve retention, particularly when multiple interventions are delivered together. However, nearly half of high-risk patients remained difficult to retain despite receiving support, with poorer outcomes observed among those who were virally unsuppressed, had depressive symptoms, or were younger

These findings suggest that integrating predictive tools into routine HIV programs could enable more efficient targeting of interventions to patients most at risk of disengagement, leading to improved retention outcomes. At the same time, more intensive and tailored strategies are needed for patients who remain at high risk despite interventions, particularly those who are virally unsuppressed, have depressive symptoms, or are younger.

## Introduction

Retention in antiretroviral therapy (ART) care remains a critical challenge to HIV program success, particularly in high-burden, resource-limited settings. Loss to follow-up (LTFU) disrupts continuity of treatment, leading to viral rebound, increased morbidity, and ongoing HIV transmission [1–3]. Across sub-Saharan Africa, retention on antiretroviral therapy declines substantially over time, with average retention estimates decreasing from approximately 79% at 6 months to 75% at 12 months, 64–74% at 24 months, and around 60% at five years, a level of attrition that undermines sustained viral suppression and weakens efforts to achieve epidemic control [4, 5]. Malawi is among the countries most affected by the HIV epidemic, with a substantial proportion of the population requiring lifelong ART. Despite substantial expansion of ART access globally, retention in Malawi remains suboptimal, with studies reporting 70–80% retention at 12–24 months, implying that approximately 20–30% of patients disengage from care during the first 2 years of ART [6, 7].

To address disengagement from care, Malawi has implemented several interventions, including the “Back-to-Care” programme, which traces individuals who miss scheduled appointments through phone calls or home visits [8]. In addition to tracing strategies, several retention-focused approaches have been adopted. These include peer-led models such as expert client (EC) support, which provide follow-up and adherence counselling; Community ART Groups (CAGs), in which small groups of patients (typically four members) rotate ART collection from the facility while providing peer adherence support; Teen Clubs, which offer age-specific peer support and adherence services for adolescents; community-based service delivery models that bring care closer to patients; and mobile health interventions, such as two-way texting (2WT), which enhance communication and engagement in care [9–11]. While these strategies have shown potential to improve retention and re-engagement, many remain reactive and resource-intensive, raising concerns about their scalability and sustainability in the context of constrained healthcare funding [12, 13]. These limitations highlight the need for more proactive and efficient approaches to support retention in care among people living with HIV.

Previous studies have explored the use of machine learning–based risk stratification for identifying patients at high risk of LTFU prior to disengagement. Supervised machine learning models have been successfully applied in multiple settings using routinely collected electronic medical record (EMR) data, demonstrating strong predictive performance across diverse populations [10–12]. Additional work incorporating unstructured clinical data has further shown the potential of machine learning to capture complex drivers of disengagement from care [14]. However, challenges remain in applying these models in routine clinical practice, including limited generalizability across settings, difficulties integrating them into existing clinical workflows, and resource constraints in low- and middle-income settings [15–17]. Furthermore, most existing studies focus primarily on predictive accuracy and do not evaluate how risk predictions can be used to guide targeted interventions or improve retention outcomes. As a result, there is limited evidence on the expected impact of machine learning–guided risk stratification for informing intervention allocation in routine ART programmes.

In an earlier phase of this study, we developed a supervised machine learning model based on Extreme Gradient Boosting (XGBoost) that demonstrated strong performance in predicting 12-month loss to follow-up among ART patients in two high-burden ART facilities in Lilongwe, Malawi [18]. Importantly, this model generated individual-level predicted probabilities of LTFU, providing a quantitative basis for identifying patients at varying levels of risk and supporting the potential for targeted intervention strategies.

However, although these models can identify patients at high risk of disengagement, there is limited evidence on how these predictions can be used to guide interventions that improve retention outcomes. Simulation-based approaches provide a way to address this gap by combining individual-level risk predictions with evidence-based estimates of intervention effects. By modelling how targeted interventions may influence retention across patients with different risk profiles, simulation enables prospective evaluation of intervention strategies before real-world implementation [19]. Such approaches are particularly valuable in settings where empirical evaluation is costly, time-consuming, and operationally complex [19, 20].

To address this gap, we used a simulation-based approach that combines individual-level predicted probabilities of LTFU generated using a machine learning model with estimates of intervention effects derived from a meta-analysis of published studies to evaluate the expected impact of targeting retention interventions to high-risk patients on ART retention outcomes in routine ART programmes. A post-simulation analysis examined differences in expected retention outcomes across patient groups, identifying subgroups that remained at high risk of disengagement despite targeted interventions.

This study aims to assess the impact of machine learning–guided targeting of retention interventions on ART retention outcomes. Specifically, the study addresses the following questions: (1) How does machine learning–guided targeting of retention interventions influence ART retention outcomes? (2) How do these effects vary across patient groups? and (3) Which subgroups remain at high risk of disengagement after targeted interventions? The remainder of the paper describes the methods, presents the results, and discusses the implications of the findings.

### Related work

Several studies in sub-Saharan Africa have demonstrated the potential of machine learning approaches to predict loss to follow-up (LTFU) among patients receiving antiretroviral therapy using routinely collected electronic medical record (EMR) data. Fahey et al. showed that ensemble machine learning models applied to EMR data in Tanzania achieved moderate predictive performance, with approximately 75% accuracy in identifying patients at risk of disengagement[15]. Similarly, Esra et al. conducted a large-scale analysis in South Africa using longitudinal EMR data and found that prior visit attendance patterns were strong predictors of treatment interruption, with models predicting a substantial proportion of missed visits [16]. In Ethiopia, Endebu et al. demonstrated that multiple machine learning algorithms can be successfully applied across different facilities to classify patients at risk of LTFU, supporting the scalability of these approaches in low-resource settings[17] Despite these advances, existing studies primarily focus on predictive accuracy and identification of risk factors, with limited attention to how such predictions can be translated into targeted intervention strategies or evaluated for their impact on retention outcomes in routine ART programmes.

## Methodology

### Study design

This study employed a patient-level Monte Carlo simulation framework to estimate ART retention under a machine learning–guided intervention targeting approach. The modelling framework integrates two distinct components: individual-level risk predictions for LTFU generated using a machine learning model and intervention effect estimates obtained through a meta-analysis conducted on published studies evaluating interventions to improve ART retention.

### Study setting

The study was conducted at two Lighthouse Trust clinics in Lilongwe, Malawi: the Lighthouse Clinic at Kamuzu Central Hospital and the Martin Preuss Centre at Bwaila Hospital. Both facilities are high-volume tertiary ART clinics operating within the national HIV programme and provide comprehensive, integrated HIV services to over 35,000 clients. The facilities implement several retention strategies, including early retention support for newly initiated ART clients through the use of expert clients. They also run a community nurse-led ART programme, in which nurses provide ART services within the community through support groups. In addition, the facilities operate a back-to-care program that traces patients who miss scheduled appointments by at least 28 days. Psychosocial support services are also provided to individuals with mental health challenges.

### Data sources

Data was obtained from two primary sources. First, routinely collected electronic medical record data from Lighthouse ART clinics provided demographic and clinical variables. Second, behavioral and psychosocial variables were obtained from the Attrition Risk Stratification Tool, which was implemented as part of routine care at Lighthouse Trust clinics from 15 September 2021 to 20 June 2024. The tool captures key risk factors associated with disengagement from care, including demographic, behavioral, and psychosocial characteristics identified in the literature and through expert input from experienced ART providers. A summary of the variables included in the tool is presented in Table 1.

**Table 1.**
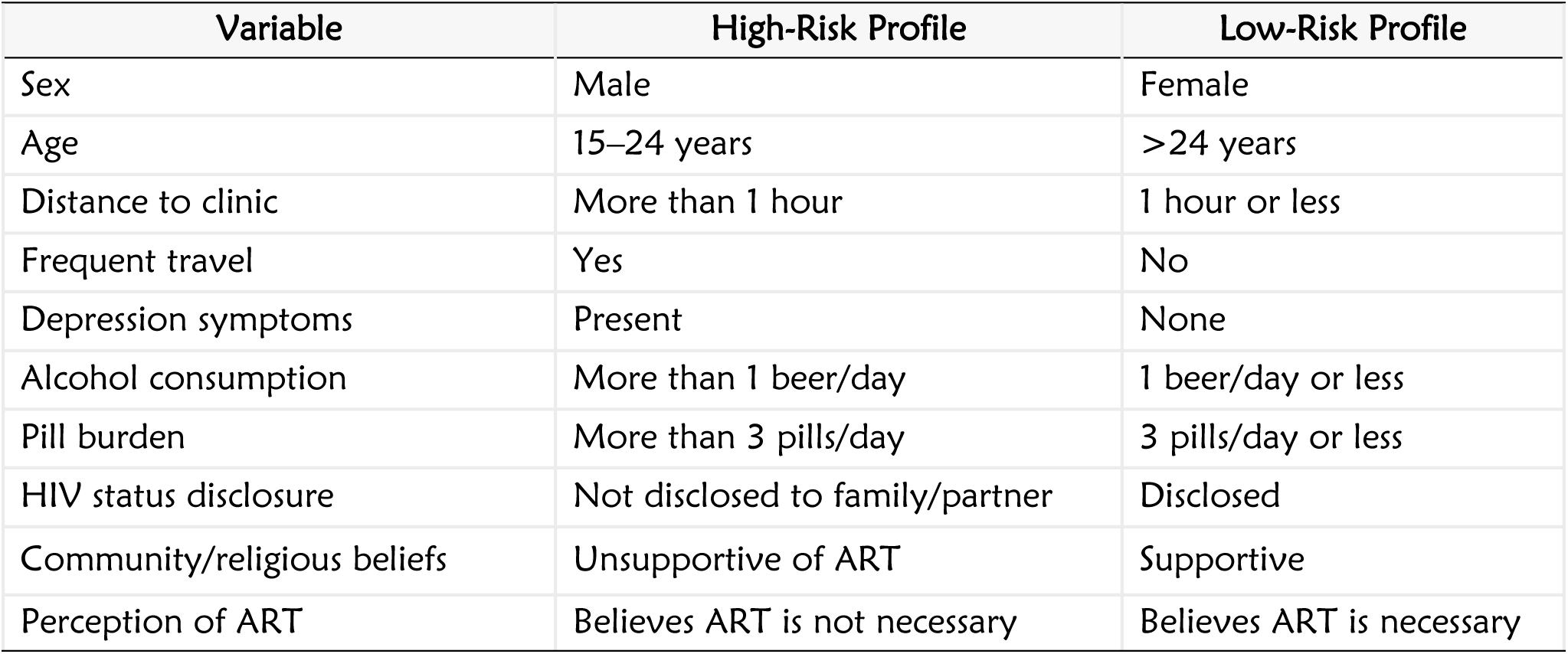
Attrition Risk Stratification tool.

| Variable | High-Risk Profile | Low-Risk Profile |
| --- | --- | --- |
| Sex | Male | Female |
| Age | 15–24 years | >24 years |
| Distance to clinic | More than 1 hour | 1 hour or less |
| Frequent travel | Yes | No |
| Depression symptoms | Present | None |
| Alcohol consumption | More than 1 beer/day | 1 beer/day or less |
| Pill burden | More than 3 pills/day | 3 pills/day or less |
| HIV status disclosure | Not disclosed to family/partner | Disclosed |
| Community/religious beliefs | Unsupportive of ART | Supportive |
| Perception of ART | Believes ART is not necessary | Believes ART is necessary |

In addition to patient-level data, intervention effect estimates were obtained through a meta-analysis conducted as part of this study, using published studies from sub-Saharan Africa that evaluated ART retention interventions. The meta-analysis produced pooled relative risk (RR) estimates, which were used to parameterize the simulation model.

### Study population

The study population consisted of 1,705 HIV-positive patients aged 15 years and older receiving ART at Lighthouse Clinic and Martin Preuss Centre in Lilongwe, Malawi. These patients represent individuals routinely accessing ART services within the study facilities.

Patients were eligible for inclusion if they were aged 15 years or older, were receiving ART at either facility, and had been screened using the Attrition Risk Stratification Tool during the study period. Records were excluded if patients had not undergone attrition risk screening or if key variables required for risk prediction and simulation modelling were missing.

### ML Prediction of LTFU risk

Individual probabilities of 12-month LTFU were generated for each patient by applying a recently developed and validated supervised machine learning model based on Extreme Gradient Boosting (XGBoost) to the study dataset. The model, trained and tested on routinely collected patient-level data, demonstrated strong predictive performance in identifying patients at risk of disengagement from care (accuracy = 0.89, precision = 0.90, recall = 0.87, F1-score = 0.88, ROC-AUC = 0.943) [18]. The predicted probabilities represent individualized estimates of disengagement from ART care over a 12-month period. Using continuous risk estimates rather than binary classifications preserves individual-level heterogeneity and supports probabilistic outcome modeling within the simulation framework, allowing for a more accurate representation of uncertainty and variability in retention outcomes [21, 22]. Patients were subsequently ranked according to their predicted probabilities of loss to follow-up, and those in the top 30% highest-risk group (approximately n = 512) were classified as high-risk. Targeted interventions were allocated exclusively to this high-risk group within the simulation framework.

### Intervention effect estimates

Effect sizes for ART retention interventions were obtained from published studies and expressed as relative risks (RR) for retention. Effect measures reported in the literature (including hazard ratios, odds ratios, risk ratios, prevalence ratios, and risk differences) were harmonised to a common metric, relative risk for retention. Conversion was implemented using standard transformation approaches, including methods appropriate for time-to-event, binary, and continuous measures, with assumptions consistent with epidemiological modelling. Pooled intervention effects were derived through a random-effects meta-analysis, producing summary relative risk estimates and corresponding 95% confidence intervals. Meta-analysis provides a quantitative approach for combining results across studies to obtain a more precise and robust estimate of intervention effects, particularly where findings vary across settings[23, 24]. A summary of the studies included in the meta-analysis and the corresponding effect estimates is provided in S1 Table. These pooled estimates were used as input parameters in the simulation model.

### Meta-analysis procedure

Following harmonisation of effect measures to relative risks for ART retention, pooled intervention effects were estimated using a random-effects meta-analysis framework. Study-specific relative risks were first transformed to the natural logarithmic scale, and corresponding standard errors were derived from reported confidence intervals. These log-transformed estimates were used as inputs for the meta-analysis.

Pooled effect estimates were calculated using the DerSimonian and Laird random-effects model, which accounts for both within-study sampling variability and between-study heterogeneity. Between-study variance was estimated using the method of moments, and inverse-variance weighting incorporating this variance component was applied to obtain pooled estimates.

Heterogeneity across studies was assessed using the Cochran Q statistic and quantified using the I² metric, providing measures of absolute and relative heterogeneity, respectively. Pooled effect estimates and their corresponding 95% confidence intervals were then back-transformed to the relative risk scale for interpretation and use in the simulation model.

### Simulation model

A patient-level Monte Carlo simulation model was constructed to combine individual-level baseline risk with intervention effect sizes in order to generate predicted retention outcomes under different scenarios. The simulation framework represents uncertainty in patient outcomes by repeatedly sampling from probability distributions at the individual level.

For each patient *i*, a baseline probability of retention was first defined using the predicted probability of LTFU obtained from the machine learning model:

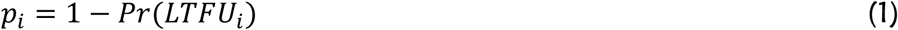

This formulation directly converts the predicted probability of LTFU into the corresponding probability of being retained in care. In this context, *p_i_* represents the expected probability that patient *i* remains in ART care over the 12-month period in the absence of additional intervention.

Intervention effects were incorporated by modifying these baseline probabilities. Each intervention was associated with a RR for retention, representing the multiplicative change in the probability of remaining in care among patients receiving the intervention.

For patients receiving more than one intervention, effects were assumed to act independently and were therefore combined multiplicatively. The combined relative risk for a patient receiving *k* interventions was calculated as:

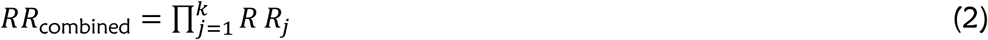

where *RR_j_* denotes the relative risk associated with intervention *j*. This formulation allows the model to represent scenarios in which multiple interventions are delivered concurrently under an assumption of independence of effects.

The adjusted probability of retention for each patient was then calculated as:

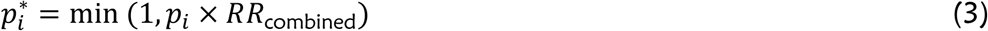

This adjustment scales the baseline probability of retention according to the expected effect of the intervention(s), while ensuring that the resulting probabilities remain within the valid range of 0 to 1.

Following this, outcomes were simulated at the individual level by sampling from a binomial distribution with *n* = 1(Bernoulli trials). For each patient, a binary outcome *Y_i_* was generated such that:

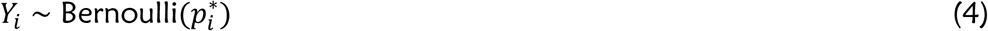

where *Y_i_* = 1 indicates that the patient is retained in care at 12 months and *Y_i_* = 0 indicates loss to follow-up.

The Monte Carlo simulation was implemented by repeating this process over 5,000 iterations for each intervention scenario. Within each iteration, retention outcomes were simulated for all patients, and overall retention was calculated. The expected retention under each scenario was defined as the mean proportion of patients retained across all simulation runs. Uncertainty was quantified using empirical 95% simulation intervals derived from the distribution of outcomes across iterations.

### Intervention scenarios and allocation

The simulation evaluated six ART retention interventions, including Expert Client support, psychosocial support, two-way text messaging, adherence clubs, community ART groups, and teen clubs. These interventions were selected based on existing evidence of effectiveness in improving ART retention[9, 25–29].

Expert Client support involves trained, stable ART patients providing peer counselling, adherence support, and follow-up to individuals at risk of disengagement [9]. In the simulation, this intervention was primarily targeted at clinically unstable patients and newly initiated clients, who are more vulnerable to early loss to follow-up. Psychosocial support includes structured counselling and mental health services that address stigma, depression, and other social barriers to care [28], and was allocated to patients with identified psychosocial or mental health needs, including those with depressive symptoms and adolescents. Two-way text messaging uses mobile phone communication to send appointment reminders, motivational messages, and enable interactive communication between patients and providers, as implemented in the Lighthouse Trust programme in Lilongwe [25], and was applied to new clients and clinically high-risk individuals to support early engagement and appointment adherence. Adherence clubs are group-based models in which clinically stable patients receive ART refills, brief clinical assessments, and peer support in a streamlined setting [27, 30], and were therefore assigned to stable patients established on ART. Community ART groups involve small groups of patients who take turns collecting medication for the group, reducing travel burden and improving peer accountability [11, 26], and were also implemented among stable patients eligible for differentiated service delivery models. Teen clubs are adolescent-focused service delivery models that combine clinical care with peer support, education, and counselling to improve engagement in care [10, 29], and were specifically provided to adolescents and young adults less than 25 years old.

Interventions were evaluated both at the overall high-risk population level and within five specific patient subgroups, including adolescents and young adults (AYA), stable clients, clinically unstable clients, Clients reporting depressive symptoms, and newly initiated ART clients. Adolescents and young adults (AYA) were defined as individuals aged <25 years. Stable Clients were defined as those with suppressed viral load, no TB or KS episodes, and aged ≥20 years. Clinically unstable patients were defined as those with unsuppressed viral load, had episode of TB, KS, advanced HIV disease, or <6 months on ART. Patients classified as depressed were those who reported experiencing depressive symptoms during routine attrition risk screening. New clients were defined as <12 months on ART, and overall high-risk population were those individuals in the top 30% of predicted risk scores. This stratified approach allowed assessment of how intervention effectiveness may vary across different patient profiles.

Each intervention was evaluated individually and, where applicable, in combination with others to reflect alternative programmatic strategies. Intervention allocation was based on alignment between patient characteristics and the intended target population of each intervention, consistent with established ART service delivery models. Patients could receive either a single intervention or multiple interventions, as defined by the intervention packages evaluated across simulation scenarios. Interventions were combined where individuals met criteria for more than one intervention category, reflecting routine programmatic practice of layered support.

### Outcome measures

The primary outcome of interest was ART retention, defined as the proportion of patients remaining in care at 12 months under each simulated intervention scenario. Secondary outcomes included the absolute change in retention relative to baseline and the relative improvement in retention under each intervention scenario.

### Sensitivity analysis

To assess the robustness of the findings, a probabilistic sensitivity analysis was conducted. Intervention effect sizes were varied within their respective 95% confidence intervals, and the simulation was repeated under these alternative parameter values. This analysis evaluated the extent to which uncertainty in intervention effectiveness influenced the estimated retention outcomes.

### Post-simulation analysis

Simulation outputs were subsequently used to classify high-risk patients into two groups based on their predicted probability of retention after intervention. Patients whose expected retention probability remained below 50% were classified as hard to retain, while those with probabilities equal to or greater than 50% were classified as responsive to existing interventions. This cutoff was selected as a pragmatic and intuitive decision boundary, representing the point at which a patient is more likely than not to be retained in care.

This post-simulation classification enabled further exploration of patient-level heterogeneity and facilitated identification of subgroups for whom additional or alternative intervention strategies may be required.

All simulations were implemented in Python, and outputs were exported to Microsoft Excel files to support transparency, reproducibility, and downstream analysis.

### Ethical Approval and Waiver of Consent

This study used routinely collected clinical and programmatic data from patients under routine care conditions. Ethical approval for use of routine data for the evaluation was granted by the Malawi National Health Sciences Research Committee (NHSRC), protocol number NHSRC #2812. A waiver of informed consent was granted for use of routinely collected program data, as approved under the data use protocol. The waiver complied with US regulations (OHRP-45CFR46.116(d)), as the retrospective evaluation posed minimal risk and did not affect participants’ rights or welfare. This study was also approved by Malawi University of Science and Technology Research Ethics Committee (MUSTREC) approval number P.02/2026/552. A comprehensive list and documentation of all ethical approvals are provided in Other_ Ethical Approval.

## Results

### General Characteristics of the study participants

Table S2 presents the general characteristics of the 1704 study participants, including demographic, clinical, behavioural, and psychosocial variables. The mean age at ART initiation was 31.6 years (SD 18.4), and 55.0% of participants were female. Overall, 95.0% of patients had suppressed viral load, and 14.6% were lost to follow-up after 12 months follow up period.

### Effect of Individual Interventions on ART Retention

Among the overall high-risk clients (n = 512), all 3 individual interventions resulted in significant improvements in ART retention compared with baseline. Baseline expected retention was 44.1% (n = 225.9). Expert Client support increased expected retention to 55.0% (n = 281.38; 95% CI: 267–296; RR = 1.25; p < 0.001), corresponding to an absolute increase of 10.9 percentage points. Psychosocial support increased retention to 52.2% (n = 267.35; 95% CI: 252–283; RR = 1.18; p < 0.001), while two-way texting increased retention to 52.7% (n = 269.72; 95% CI: 254–285; RR = 1.19; p < 0.001), representing absolute improvements of 8.1 and 8.6 percentage points, respectively. Across all interventions, the 95% confidence intervals did not overlap with baseline estimates, indicating consistent improvements in retention across simulated scenarios (Fig 1).

**Fig 1.**
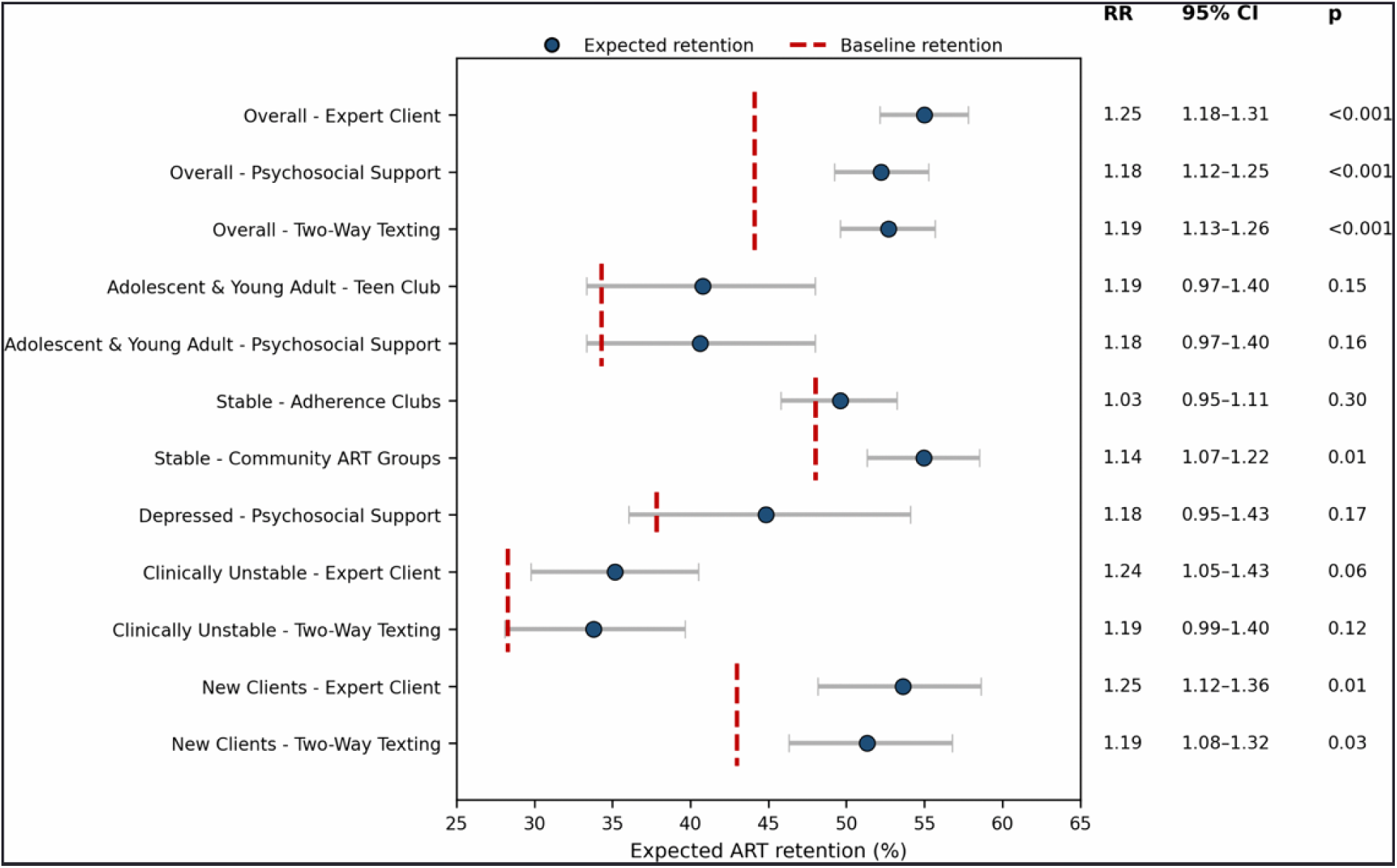
Expected ART retention under individual targeted interventions. Points represent mean expected ART retention (%) under each intervention, with horizontal lines indicating 95% uncertainty intervals derived from Monte Carlo simulations. Red dashed vertical lines denote baseline retention levels for each corresponding patient subpopulation. The table on the right shows estimated risk ratios 95% confidence intervals (CI), and p-values comparing each intervention scenario to its respective baseline. Statistically significant improvements (p < 0.05) were observed for several interventions.

Subgroup analyses revealed variability in the effectiveness of retention interventions across different patient populations. Among adolescents and young adults (n = 75), baseline retention was 34.3%. Both Teen Club participation and psychosocial support were associated with increases in expected retention to 40.8% (RR = 1.19; p = 0.15) and 40.6% (RR = 1.18; p = 0.16), respectively. However, these improvements did not reach statistical significance. In clinically stable patients (n = 417), baseline retention was higher at 48.0%. Adherence Clubs resulted in a minimal increase in retention to 49.6% (RR = 1.03; p = 0.30), indicating no significant effect. In contrast, Community ART Groups significantly improved retention to 54.9% (RR = 1.14; p = 0.01). Among patients with depressive symptoms (n = 61), baseline retention was 37.9%. Psychosocial support increased retention to 44.8% (RR = 1.18; p = 0.17), although this improvement was not statistically significant.

Among clinically unstable patients (n = 121), baseline retention was 28.3%. Expert Client support increased retention to 35.2% (RR = 1.24; p = 0.06), while two-way texting increased retention to 33.8% (RR = 1.19; p = 0.12). Although both interventions showed positive effects, neither reached statistical significance. Among newly initiated ART patients (n = 162), baseline retention was 43.0%. Expert Client support increased retention to 53.6% (95% CI: 78–95 retained; RR = 1.25; p = 0.01), while two-way texting increased retention to 51.3% (95% CI: 75–92 retained; RR = 1.19; p = 0.03). Both interventions resulted in statistically significant improvements in this subgroup. Overall, intervention effectiveness varied across subpopulations, with stronger and statistically significant effects observed among newly initiated patients and among those receiving Community ART Group services, and more limited effects among adolescents, patients with depression, and clinically unstable patients. Absolute numbers underlying these estimates are provided in S3 Table.

### Effects of combined intervention packages on ART retention

As shown in Table 2, in the overall population of high-risk patients (n = 512), the combined intervention package consisting of Expert Client support, psychosocial support, and two-way texting resulted in substantial improvements in ART retention compared with baseline. Baseline retention was 44.1% (≈226 patients), and the combined intervention increased expected retention to 64.0% (≈328 patients; RR = 1.45), representing an absolute improvement of 19.9 percentage points. The 95% simulation interval ranged from 315 to 341 retained patients, indicating consistently higher retention across 5,000 simulation iterations.

**Table 2.** Expected ART retention under combined intervention packages by patient subpopulation.

| Subpopulation | Interventions | Eligible Patients | Baseline Retained (n) | Baseline Retention (%) | Expected Retained (n) | Expected Retained (%) | 95% CI | RR | p-Value |
| --- | --- | --- | --- | --- | --- | --- | --- | --- | --- |
| Overall | EC, 2WT & PSS | 512 | 225.9 | 44.1 | 327.6 | 63.9 | 315–341 | 1.45 | <0.001 |
| Teen | Teen Club & PSS | 75 | 25.73 | 34.31 | 34.17 | 45.57 | 29–40 | 1.33 | 0.03 |
| Stable | Adherence Clubs, CAGS | 417 | 200.32 | 48.04 | 236.59 | 56.74 | 222–251 | 1.18 | <0.001 |
| Depressed | PSS | 61 | 23.09 | 37.85 | 27.31 | 44.77 | 22–33 | 1.18 | 0.18 |
| Clinically unstable | EC & 2WT | 121 | 34.25 | 28.31 | 47.32 | 39.11 | 41–54 | 1.38 | 0.01 |
| New Clients | EC & 2WT | 162 | 69.62 | 42.97 | 95.51 | 58.96 | 88–103 | 1.37 | <0.001 |

Similar patterns of improvement were observed across subpopulations, although the magnitude of effects varied according to baseline retention levels. Among adolescents and young adults (n = 75), baseline retention was 34.3% (≈26 patients), adherence and teen clubs interventions combined increased retention to 45.6% (≈34 patients; RR = 1.33), corresponding to an absolute improvement of 11.3 percentage points (95% simulation interval: 29–40 patients). Among clinically stable patients (n = 417), baseline retention was 48.0% (≈200 patients), and the combination of Adherence Clubs and Community ART Groups increased retention to 56.8% (≈237 patients; RR = 1.18), representing an absolute improvement of 8.7 percentage points (95% simulation interval: 222–251 patients).

Among clinically unstable patients (n = 121), baseline retention was 28.3% (≈34 patients), and the combined intervention of Expert Client support and two-way texting increased retention to 39.1% (≈47 patients; RR = 1.38), corresponding to an absolute improvement of 10.8 percentage points (95% simulation interval: 41–54 patients). Among newly initiated ART patients (n = 162), baseline retention was 43.0% (≈70 patients), increasing to 58.9% (≈95 patients; RR = 1.37), representing an absolute gain of 15.9 percentage points (95% simulation interval: 87–104 patients).

### Sensitivity analysis results

In the Probabilistic sensitivity analysis, Across all subpopulations and intervention scenarios, expected retention consistently exceeded baseline levels, and relative risks for retention remained greater than one. The magnitude and ordering of intervention effects were preserved under sensitivity variation, with larger absolute gains observed among subgroups with lower baseline retention, including high-risk clients, clinically unstable patients, and newly initiated ART patients. Simulation-derived uncertainty intervals widened appropriately in smaller subgroups but did not materially alter interpretation of the results. Overall, the sensitivity analysis confirms that the main simulation findings are stable and not driven by narrow or optimistic assumptions regarding intervention effectiveness.

### Post-intervention retention outcomes within the high-risk cohort

Within the high-risk cohort (n = 512), patients were classified based on their expected probability of retention following assignment of all eligible interventions. Patients with expected retention below 50% were categorized as *hard to retain*, while those with higher probabilities were classified as *improved retention*. Overall, 236 patients (46%) were classified as hard to retain, and 276 (54%) demonstrated improved retention (Table 3).

**Table 3.** Post-intervention retention outcomes within the high-risk cohort.

| Metric | Improved retention | Remained hard to retain |
| --- | --- | --- |
| Number of patients (n) | 276 | 236 |
| Mean age (years) | 33.7 | 33.7 |
| Mean duration on ART (months) | 20.3 | 19.7 |
| Baseline retention (%) | 69 | 15 |
| Expected retention (%) | 88 | 20 |
| Absolute change (%) | 19 | 5 |
| Mean number of interventions | 2.54 | 2.18 |

The two groups were comparable with respect to demographic characteristics. Mean age was identical in both groups (33.7 years), and duration on ART was similar, with 20.3 months among patients with improved retention and 19.7 months among those who remained hard to retain. Despite these similarities, substantial differences were observed in both baseline and post-intervention retention outcomes.

Patients with improved retention had a markedly higher baseline retention (69%) compared with those who remained hard to retain (15%). Following intervention assignment, expected retention increased to 88% among patients with improved retention, whereas retention among the hard-to-retain subgroup remained low at 20%. Although patients with improved retention received slightly more interventions on average (2.54 vs 2.18), the hard-to-retain subgroup continued to exhibit substantially lower expected retention despite multiple interventions, indicating persistent vulnerability to disengagement from care.

### Post-intervention subgroup patterns by age, viral load suppression, and depression

Post-intervention retention patterns across age group, viral load suppression status, and depression status within the high-risk cohort are visualized in Fig 2, with detailed subgroup estimates provided in S4 Table. Substantial heterogeneity in retention outcomes was observed across subgroup combinations. Among patients aged <25 years, retention outcomes varied markedly by viral load suppression and depression status. Patients who were virally suppressed and not depressed (n = 36) had an expected retention of 63%, with 38.9% classified as hard to retain. In contrast, suppressed patients with depression (n = 7) had lower expected retention (46%) and a higher proportion classified as hard to retain (71.4%). Among unsuppressed patients, expected retention was 34% in those without depression and only 4% among those with depression, with all patients in this latter subgroup classified as hard to retain.

**Fig 2.**
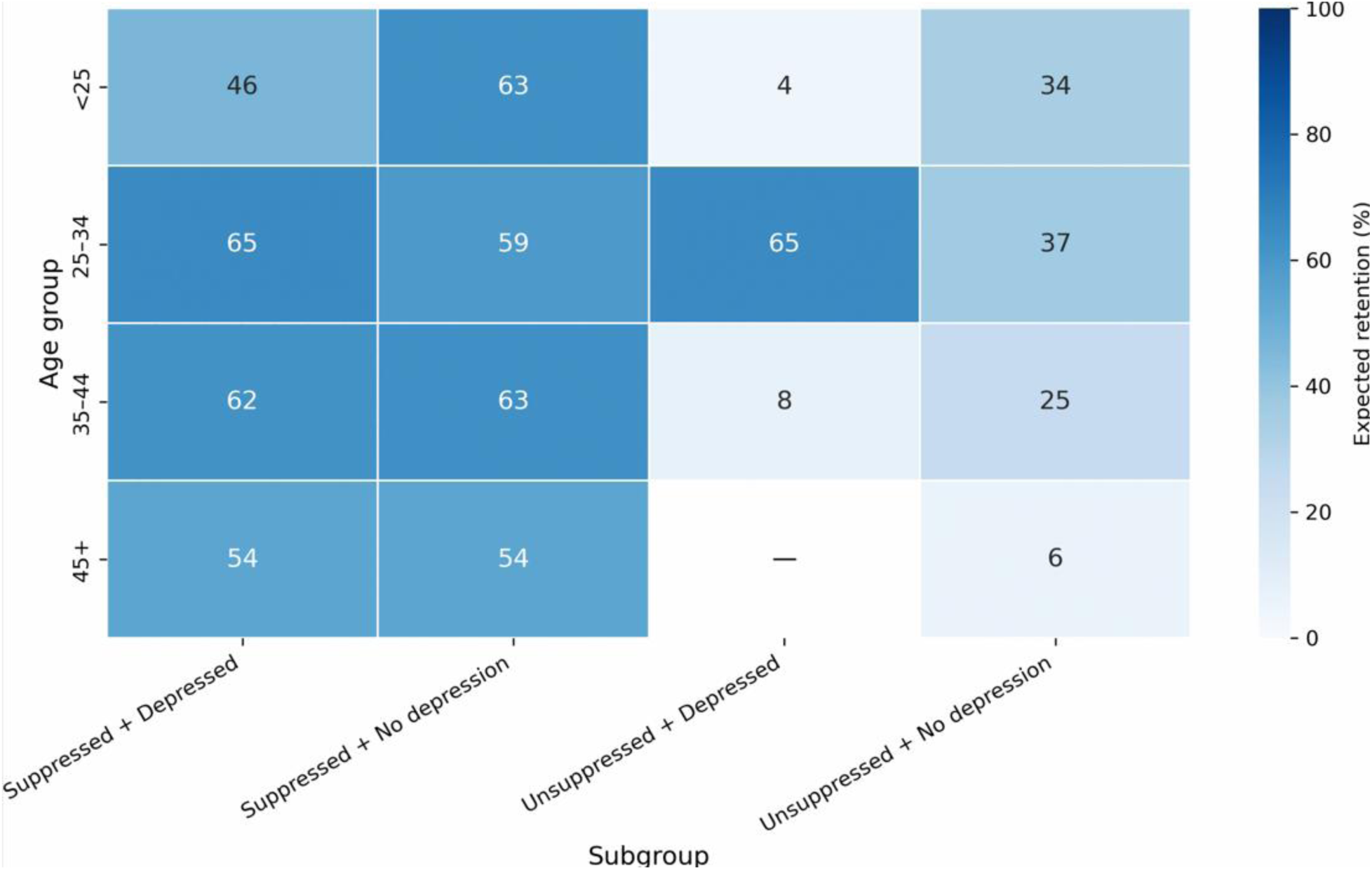
Heatmap of post-intervention ART retention across subgroups within the high-risk cohort. Each cell represents expected retention (%) following intervention across subgroups defined by age, viral load suppression, and depression status. Lower retention is observed among unsuppressed patients, particularly those with depression, indicating substantial heterogeneity in intervention response.

Among patients aged 25–34 years, expected retention among virally suppressed individuals ranged from 59% to 65%, with 33.3%–44.0% classified as hard to retain. In contrast, unsuppressed patients without depression (n = 16) had lower expected retention (37%) and a higher proportion classified as hard to retain (68.8%). Notably, suppressed patients with depression (n = 27) received the highest mean number of interventions (4.15) and exhibited the lowest proportion classified as hard to retain (33.3%).

In the 35–44-year age group, virally suppressed patients maintained expected retention above 60%, with fewer than 45% classified as hard to retain. In contrast, unsuppressed patients showed substantially lower retention, with expected retention of 25% among those without depression and 8% among those with depression, and high proportions classified as hard to retain (87.5% and 100%, respectively).

Among patients aged 45 years and older, expected retention among virally suppressed individuals was moderate (54%), with approximately half classified as hard to retain. All unsuppressed patients in this age group (n = 4) were classified as hard to retain, with very low expected retention (6%). No patients were observed in the unsuppressed and depressed subgroup in this age category.

## Discussion

This study demonstrates that machine learning–guided targeting of retention interventions has substantial potential to improve ART retention among patients at high risk of LTFU. By integrating individual-level risk predictions with intervention effect estimates in a Monte Carlo simulation framework, the analysis provides quantitative evidence that targeted, risk-informed strategies can meaningfully enhance retention outcomes compared with untargeted approaches. The findings extend beyond predictive modelling by illustrating how risk stratification can be operationalized to inform intervention allocation and improve programme performance.

Across all simulated scenarios, retention improved consistently when interventions were targeted to high-risk patients. Individual interventions produced moderate gains, while combined intervention packages generated the largest improvements, increasing retention by approximately 20 percentage points in the overall high-risk cohort. These findings suggest that no single intervention is sufficient to address the multifactorial drivers of disengagement from ART care. Instead, layered approaches that simultaneously address behavioral, psychosocial, and structural barriers appear to be more effective. This aligns with existing evidence showing that disengagement from HIV care is rarely driven by a single factor but rather by the interaction of competing life demands, stigma, mental health challenges, economic, and health system barriers[31–33].

The observed intervention effects are consistent with prior studies in Malawi and similar settings. Peer-led approaches such as Expert Client support likely improve retention by enhancing trust, providing practical adherence support, and facilitating navigation of care services [34]. Similarly, two-way text messaging and psychosocial interventions can improve engagement by addressing communication gaps and reducing psychological barriers to care [25, 35]. The relatively larger gains observed when these interventions were combined suggest synergistic effects, where multiple barriers are addressed simultaneously. This reinforces the importance of integrated service delivery models within ART programmes, particularly for patients with complex needs.

These findings highlight substantial heterogeneity in intervention response, even among patients classified as high risk. Although overall retention improved, nearly half of high-risk patients remained hard to retain even after receiving multiple interventions. These individuals were disproportionately characterized by viral non-suppression, depressive symptoms, and, in some cases, younger age. The persistence of low retention in this subgroup likely reflects deeper structural and clinical challenges that are not adequately addressed by standard interventions. For example, untreated depression can directly impair adherence and clinic attendance[36], while ongoing viremia may indicate underlying adherence difficulties, treatment fatigue, or social instability[37, 38] These findings suggest that current differentiated service delivery strategies, while effective for many patients, may not be sufficient for individuals with compounded vulnerabilities.

The subgroup analyses further highlight important patterns in intervention effectiveness. Patients who were newly initiated on ART showed comparatively larger improvements, suggesting that early engagement represents a critical window for intervention. In contrast, more modest gains among clinically unstable patients and those with depressive symptoms indicate that additional or more intensive support strategies may be required for these groups. Together, these findings underscore the importance of tailoring interventions not only based on predicted risk but also on underlying patient characteristics and needs [32, 33].

From a programmatic perspective, these results support a shift from reactive, broad-based retention strategies toward proactive, data-driven approaches. In many settings, interventions such as tracing or adherence support are deployed after patients have already disengaged from care. In contrast, the approach evaluated in this study enables earlier identification of high-risk individuals and prioritization of interventions before disengagement occurs. This has important implications for resource allocation in resource-limited settings, where maximizing the impact of available interventions is critical. Targeting high-risk patients with more intensive, combined interventions may yield greater improvements in retention than distributing resources uniformly across all patients[39].

In addition, the study provides a practical framework for integrating machine learning into routine ART programmes. By generating individual-level risk probabilities, the model supports continuous risk stratification that can be updated as patient data evolve. When linked to predefined intervention packages, such systems could be embedded within electronic medical record platforms to support real-time decision-making by healthcare providers. This represents a feasible pathway for translating predictive analytics into actionable clinical and programmatic strategies in low-resource settings.

This study also makes a methodological contribution by explicitly linking predictive modelling with intervention impact through simulation. While many studies have demonstrated the ability of machine learning models to predict loss to follow-up, few have evaluated how these predictions can be used to improve patient outcomes. By combining predictive outputs with meta-analytic estimates of intervention effectiveness, the current approach bridges this gap and provides a forward-looking assessment of potential programme impact. This type of simulation-based evidence is particularly valuable in contexts where large-scale experimental evaluations are costly or operationally challenging [19, 20].

This study had several limitations that should be considered when interpreting the findings: First, the analysis is based on simulation rather than observed intervention outcomes and therefore estimates potential effects under modeled conditions rather than fully capturing real-world effectiveness. Second, the simulation relies on simplifying assumptions, including that intervention effects combine independently and multiplicatively, which may not fully capture potential interactions or synergies between interventions. Third, the intervention effect sizes were derived primarily from studies conducted in sub-Saharan Africa, including some from Lilongwe where this study was conducted; however, variation in study populations, implementation conditions, and settings may still limit their direct applicability to the broader Malawian context.

Future research should focus on validating these findings in real-world implementation settings. Prospective studies are needed to evaluate the effectiveness, feasibility, and cost-effectiveness of machine learning–guided intervention targeting within ART programmes. In addition, further work is required to develop and test more intensive or specialized interventions for patients who remain at high risk despite receiving standard support. Refinement of modelling approaches to incorporate dynamic patient trajectories and interaction effects between interventions would also strengthen the evidence base.

## Conclusion

This study demonstrates that integrating machine learning–based risk stratification with targeted allocation of evidence-based retention interventions can substantially improve ART retention, particularly among patients at high risk of loss to follow-up. Simulation results showed consistent gains across intervention scenarios, with the greatest improvements observed under combined intervention strategies.

The use of patient-level predicted probabilities allowed for a more detailed assessment of individual risk and variation in outcomes. Notably, patients who were virally unsuppressed, had depression, and younger individuals remained at high risk of disengagement despite receiving multiple interventions. These findings highlight important limitations of current retention strategies and underscore the need for more intensive and tailored approaches for these high-risk subgroups.

This study makes several important contributions. First, it moves beyond prediction-focused machine learning analyses by explicitly linking individual-level risk predictions to the targeting of retention interventions. Second, by integrating individual-level predicted probabilities of loss to follow-up with a simulation-based framework, it provides a novel approach for prospectively evaluating the impact of targeted intervention strategies on ART retention outcomes before real-world implementation. Third, it identifies hidden patterns of residual disengagement risk that are not apparent in standard analyses, revealing patient subgroups that continue to exhibit high predicted risk of loss to follow-up or limited improvement in retention outcomes despite receiving interventions, thereby providing critical insights for refining and optimizing retention strategies. Overall, these findings provide practical evidence on how predictive analytics can be operationalized to support more efficient, proactive, and patient-centered allocation of resources within ART programmes. Future work should prioritize real-world implementation and integration of machine learning–guided intervention targeting within routine ART programmes to evaluate feasibility, scalability, and impact on long-term retention outcomes.

## Data Availability

The dataset supporting the conclusions of this study is included within the article and its supplementary files (S1 Data).

## Acknowledgments

We acknowledge the management of Lighthouse Trust for their support and guidance. We also thank Lighthouse Trust personnel who supported data collection and data management.

## Author contributions

Writing – original draft: AT,

Formal analysis: AT,

Conceptualization: AT, BK

Methodology: AT, BK

Investigation: AT, BK, ER, JH, LG, EV

Data curation: AT, EV

Validation: AT

Supervision: AT, BK, LG, JH, ER

Project administration: AT, ER, BK

Resources: AT, ER

Writing – review & editing: All authors

Visualisation: AT

## Supporting Information

**S1 Table.** Studies included in the meta-analysis and corresponding intervention effect estimates for ART retention. The table presents study characteristics, populations, intervention types, reported effect measures, and harmonized relative risks for retention (RRret) with 95% confidence intervals used in the simulation model.

**Table S2.** Baseline characteristics of study participants, including demographic, clinical, behavioral, and treatment-related factors, summarized as mean (SD) or frequency (%), among individuals on ART, with loss to follow-up (LTFU) as the outcome of interest (N = 1,704).

| Variable | Category | Value |
| --- | --- | --- |
| Months on ART | Mean (SD) | 25.6 (19.4) |
| Age at ART Initiation | Mean (SD) | 31.6 (18.4) |
| Gender | Female | 938 (55.0%) |
|  | Male | 766 (45.0%) |
| Time to Clinic | ≤1 hour | 887 (52.1%) |
|  | >1 hour | 817 (47.9%) |
| Frequent Travel | No | 1,389 (81.5%) |
|  | Yes | 315 (18.5%) |
| Feeling Depressed | No | 1,460 (85.7%) |
|  | Yes | 244 (14.3%) |
| Alcohol Uptake | ≤1 beer/day | 1,483 (87.0%) |
|  | >1 beer/day | 221 (13.0%) |
| Pill Burden | ≤3 pills/day | 1,200 (70.4%) |
|  | >3 pills/day | 504 (29.6%) |
| External Influence | Leaders support ART | 1,676 (98.4%) |
|  | Leaders against ART | 28 (1.6%) |
| Disclosure | Yes | 1,565 (91.8%) |
|  | No | 139 (8.2%) |
| Perception of ART | Believes ART helps | 1,698 (99.6%) |
|  | Just well without ART | 6 (0.4%) |
| HIV Stage | Stage 1 & 2 | 1,293 (75.9%) |
|  | Stage 3 & 4 | 411 (24.1%) |
| Last Viral load status | Suppressed | 1620 (95.0%) |
|  | Not suppressed | 85 (5.0%) |
| Hypertensive | No | 1243 (72.9%) |
|  | Yes | 462 (27.1%) |
| TB episode | No | 1528 (89.6%) |
|  | Yes | 177 (10.4%) |
| LTFU (target) | No | 1,456 (85.4%) |
|  | Yes | 248 (14.6%) |

**S3 Table.** Expected ART retention under individual interventions by patient subpopulation.

| Population | Intervention | Eligible Patients | Baseline Retained (n) | Baseline Retention (%) | Expected Retained (n) | Expected Retained% | 95% CI | RR | p_value |
| --- | --- | --- | --- | --- | --- | --- | --- | --- | --- |
| Overall | Expert Client | 512 | 225.9 | 44.13 | 281.38 | 55 | 267–296 | 1.25 | <0.001 |
| Overall | Psychosocial Support | 512 | 225.9 | 44.13 | 267.35 | 52.2 | 252–283 | 1.18 | <0.001 |
| Overall | Two-way Texting | 512 | 225.9 | 44.13 | 269.72 | 52.7 | 254–285 | 1.19 | <0.001 |
| Adolescents & young adult | Teen Club | 75 | 25.73 | 34.31 | 30.6 | 40.8 | 25–36 | 1.19 | 0.15 |
| Adolescents & young adult | Psychosocial Support | 75 | 25.73 | 34.31 | 30.46 | 40.61 | 25–36 | 1.18 | 0.16 |
| Stable | Adherence Clubs | 417 | 200.32 | 48.04 | 206.81 | 49.59 | 191–222 | 1.03 | 0.3 |
| Stable | Community ART Groups | 417 | 200.32 | 48.04 | 229.16 | 54.95 | 214–244 | 1.14 | 0.01 |
| Depressed | Psychosocial Support | 61 | 23.09 | 37.85 | 27.35 | 44.84 | 22–33 | 1.18 | 0.17 |
| Clinically unstable | Expert Client | 121 | 34.25 | 28.31 | 42.55 | 35.16 | 36–49 | 1.24 | 0.06 |
| Clinically unstable | Two-way Texting | 121 | 34.25 | 28.31 | 40.87 | 33.78 | 34–48 | 1.19 | 0.12 |
| New Clients | Expert Client | 162 | 69.62 | 42.97 | 86.85 | 53.61 | 78–95 | 1.25 | 0.01 |
| New Clients | Two-way Texting | 162 | 69.62 | 42.97 | 83.16 | 51.33 | 75–92 | 1.19 | 0.03 |

This table presents absolute and percentage ART retention outcomes under individual intervention scenarios for each patient subpopulation. Baseline and expected retained counts are reported alongside percentage retention and 95% uncertainty intervals derived from Monte Carlo simulations. Risk ratios (RR), 95% confidence intervals (CI), and p-values compare each intervention scenario to the corresponding baseline.

**S4 Table.** Post-intervention ART retention outcomes across subgroups defined by age, viral load suppression, and depression status within the high-risk cohort.

| Age group | Viral load | Depression | Patients (n) | Baseline retention (%) | Expected retention (%) | Hard-to-retain (%) | Mean interventions |
| --- | --- | --- | --- | --- | --- | --- | --- |
| <25 | Suppressed | No | 36 | 45 | 63 | 38.9 | 3.5 |
| <25 | Suppressed | Yes | 7 | 32 | 46 | 71.4 | 3.86 |
| <25 | Unsuppressed | No | 9 | 20 | 34 | 66.7 | 3 |
| <25 | Unsuppressed | Yes | 3 | 2 | 4 | 100 | 3 |
| 25–34 | Suppressed | No | 184 | 47 | 59 | 44 | 2.28 |
| 25–34 | Suppressed | Yes | 27 | 45 | 65 | 33.3 | 4.15 |
| 25–34 | Unsuppressed | No | 16 | 30 | 37 | 68.8 | 1.25 |
| 25–34 | Unsuppressed | Yes | 3 | 45 | 65 | 33.3 | 3 |
| 35–44 | Suppressed | No | 135 | 51 | 63 | 38.5 | 1.94 |
| 35–44 | Suppressed | Yes | 14 | 41 | 62 | 42.9 | 4.07 |
| 35–44 | Unsuppressed | No | 16 | 20 | 25 | 87.5 | 1 |
| 35–44 | Unsuppressed | Yes | 3 | 5 | 8 | 100 | 3 |
| 45+ | Suppressed | No | 51 | 43 | 54 | 49 | 2.02 |
| 45+ | Suppressed | Yes | 4 | 34 | 54 | 50 | 4 |
| 45+ | Unsuppressed | No | 4 | 5 | 6 | 100 | 1 |
| 45+ | Unsuppressed | Yes | 0 | — | — | — | — |

